# On Statistical Power for Case-Control Host Genomic Studies of COVID-19

**DOI:** 10.1101/2020.08.26.20182840

**Authors:** Yu-Chung Lin, Jennifer D. Brooks, Shelley B. Bull, France Gagnon, Celia M.T. Greenwood, Rayjean J. Hung, Jerald Lawless, Andrew Paterson, Lei Sun, Lisa Strug

## Abstract

The identification of genetic variation that directly impacts infection susceptibility and disease severity of COVID-19 is an important step towards risk stratification, personalized treatment plans, therapeutic and vaccine development and deployment. Given the importance of study design in infectious disease genetic epidemiology, we use simulation and draw on current estimates of exposure, infectivity and test accuracy of COVID-19 to demonstrate the feasibility of detecting host genetic factors associated with susceptibility and severity with published COVID-19 study designs. We demonstrate why studying susceptibility to SARS-CoV-2 infection could be futile at the early stages of the pandemic. Our insights can aid in the interpretation of genetic findings emerging in the literature and guide the design of future host genetic studies.

## Background

Variability in susceptibility and disease severity in response to viral exposure is a hallmark of infectious diseases [1]. This variation is partially due to a complex interaction between the pathogen and the host genome, and it is impacted directly or indirectly by factors specific to both the pathogen (e.g. viral load, viral genotype) and the host (e.g. sex, pre-existing medical conditions, race) [2]. Identifying host genetic factors responsible for this variation can pinpoint susceptible populations, inform personalized treatment, and guide vaccine development [3]. SARS-CoV-2, the virus that causes Coronavirus Disease 2019 (COVID-19), appears to be no exception [4]. By Spring 2020, several international research groups [5] were proposing or implementing studies to identify the host genetic factors that influence i) *susceptibility to infection* with SARS-CoV-2 [5] and ii) *disease severity* of COVID-19 once infected [5]. These include sequencing and array-based studies that investigate rare [6] and common [5,7] genetic variants.

Numerous case-control studies of varied designs have emerged to better understand the host genetic factors associated with susceptibility and severity of COVID-19. Due to the fast-paced nature of the pandemic, one major challenge is how to best utilize existing resources while accounting for the limitations of the available phenotype and genotype data. The study designs proposed to date include prospective case-control recruitment designs where array-based genotyping or sequencing is planned [6,8], retrospective designs using existing genotyped cohorts with concurrent phenotyping for COVID-19 [9,10], and hybrid designs using prospectively collected cases and population controls with genotype data readily available [7]. Each design has implications for how the susceptibility and disease severity phenotypes are defined [11] and may have associated limitations that are not surmountable given the current state of the pandemic.

The field of infectious disease genetic epidemiology has long described the unique and complex implications that a requirement of exposure to the infectious agent imposes on study design [12]. Here we draw on current estimates of exposure, infectivity and test accuracy of COVID-19 to demonstrate the feasibility of detecting host genetic factors associated with COVID-19 susceptibility and severity with current study designs. We demonstrate why studying susceptibility to SARS-CoV-2 infection could be futile at certain stages of the pandemic. Our insights can aid in the interpretation of genetic findings emerging from the literature and guide the design of future host genetic studies.

### Identifying Host Genetic Factors of Susceptibility to Infection with SARS-CoV-2

An important objective of host genetic studies is to determine whether there are rare or common genetic variants that modify the risk of SARS-CoV-2 infection upon exposure. Examples of the existence of such variants for other infectious diseases are sparse in the literature [3], although for SARS-CoV-2, one recent study estimated 50% heritability for infection susceptibility [4]. Case-control studies to identify such genetic contributors to SARS-CoV-2 infection have, for practical reasons, defined *cases* as individuals with laboratory confirmed SARS-CoV-2 infection and *controls* as those who tested negative [11,13]. Given low population exposure rates at the early stage of the pandemic [14], the lack of data on an individual’s exposure to SARS-CoV-2 [3] and high infectivity [15], many of these “controls” will be misclassified since it is not certain whether they were actually exposed to the pathogen, and would have been infected had they been exposed. Imperfect test accuracy also contributes to misclassification. For example, concerns have been raised about false negative Reverse Transcription Polymerase Chain Reaction (RT-PCR) tests in patients showing apparent COVID-19 illness; sometimes up to 40% of samples were deemed falsely negative [16]. False positive tests, although less prevalent due to the high specificity of RT-PCR tests [17], lead to misclassification of cases, which can also be problematic if disease prevalence is low. The result of misclassification of either cases or controls is reduced statistical power for detecting common and rare genetic variants associated with susceptibility to infection with SARS-CoV-2 (Figure 1).

**Figure 1:**
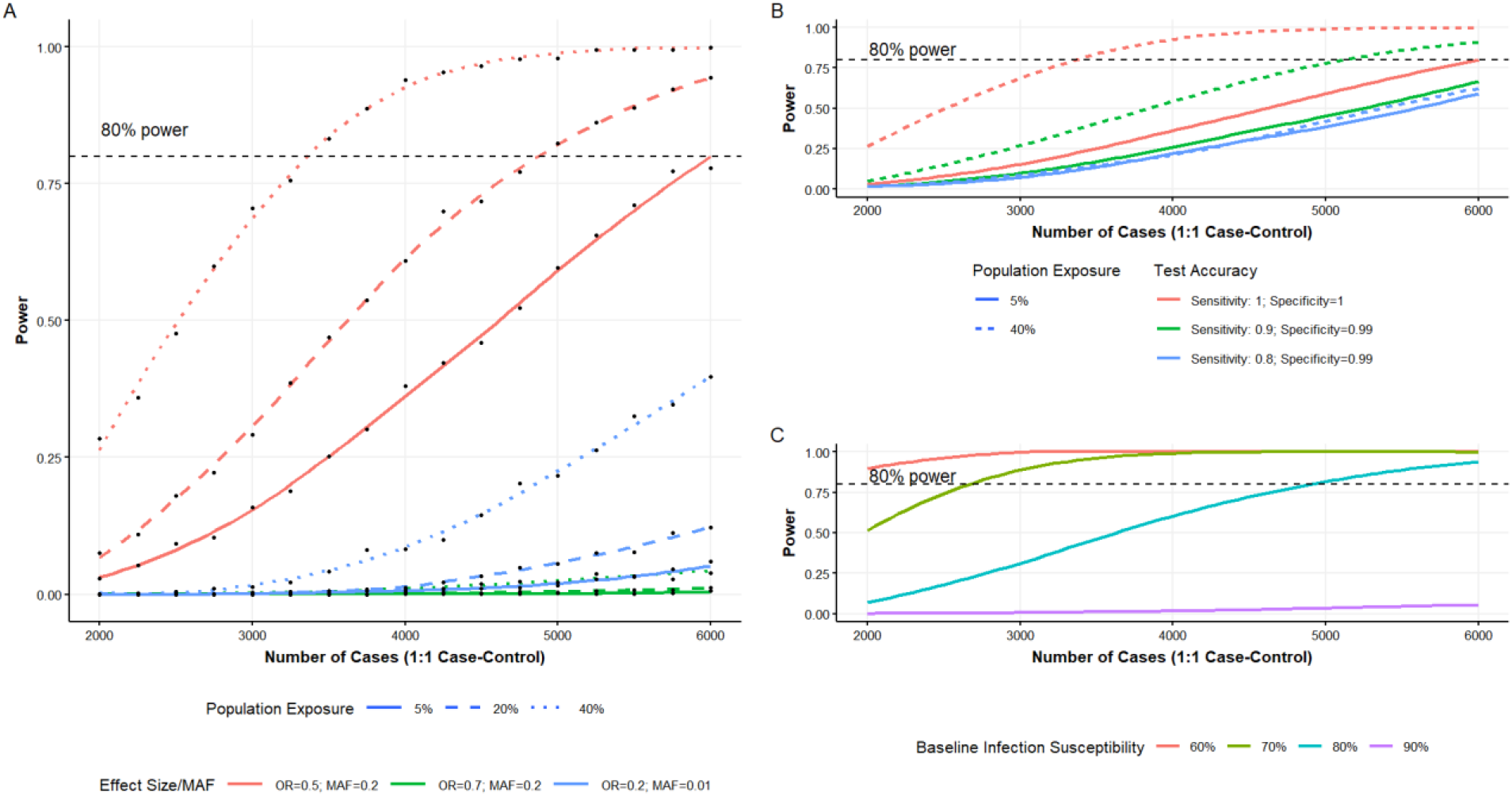
Statistical power to detect association between a genetic variant and infection susceptibility at the genome-wide significance level (5e-8) [22]. A 1:1 case-control study design was used for all parameter settings. Reported effect sizes are on the odds ratio (OR) scale, parameterized as log-additive for each additional protective allele. A) Assuming perfect test accuracy and baseline infection susceptibility at 80% based on recent estimates [11], there is low statistical power to detect true associations when there is either low population-level exposure to SARS-CoV-2 or moderate genetic (protective) effect sizes (OR=0.7). Detecting rare variants (MAF=0.01) remains challenging even with a much larger protective effect size (OR=0.2); B) Reducing sensitivity for testing SARS-CoV-2 infection not only reduces statistical power but also negates gains that result from increasing population exposure. C) Assuming 20% population exposure rate seen in the hardest-hit regions, baseline infection susceptibility, in the absence of the contributing protective genetic allele, can also severely impact power. Higher infection susceptibility (i.e. higher infectivity) can diminish any chance of detecting true signals with currently available sample sizes. Lower population exposure will further dampen statistical power as seen in Figure 1A. MAF=minor allele frequency.

We used simulations to demonstrate the factors that impact statistical power to identify genetic variants associated with SARS-CoV-2 infection susceptibility. It is assumed that controls are selected from persons testing negative for the virus. In addition to sample size, effect size of a variant (odds ratio; OR) and its minor allele frequency (MAF), critical factors include rates of population-level exposure, test accuracy and baseline infection susceptibility (Figure 1 and Supplement A). We assume that the majority of the population will be infected when exposed, except those who carry protective alleles [18, 19]. Therefore, the power calculation is estimated based on a range of ORs less than 1, without loss of generality.

Figure 1A demonstrates that under perfect test accuracy, not only is it difficult to detect rare variants with very large protective effects (OR=0.2; MAF=0.01; blue), it is extremely challenging to detect common variants with moderate protective effects (OR=0.7; MAF=0.2; green) using sample sizes reported in existing genetic studies [5]. Moreover, low population-level exposure (5%; solid line Figure 1A) reduces statistical power, as many of the “controls” would have been infected if exposed to SARS-CoV-2, contributing to higher misclassification rates. Given low population exposure rates even in the hardest-hit regions [20], our chances of detecting a true association with infection susceptibility remain slim. The curation of better-defined controls through careful selection of those with a high probability of exposure to SARS-CoV-2 (such as health care workers or household members of positive cases), which is analogous to an increasing population-level exposure, leads to improved power to detect associated genetic variants.

Figure 1B demonstrates that false negative tests caused by low sensitivity of SARS-CoV-2 infection testing [16] can dramatically reduce statistical power. Increasing the number of false negative tests (sensitivity=0.9, green; sensitivity=0.8, blue) leads to much reduced study power even when the genetic variant of interest is a common variant with large genetic (protective) effect (OR=0.5, MAF=0.2; Figure 1B). Increasing population-level exposure or using better-defined controls can improve power (Figure 1A), but these power gains are no longer guaranteed when coupled with an inaccurate, low-sensitivity test for SARS-CoV-2 infection. Increasing population exposure does not result in higher statistical power when sensitivity=0.8 (blue curve; Figure 1B) since the test misclassifies many cases as controls and offsets any power gain that would be realized by carefully selecting exposed individuals as controls. With lower test sensitivity (sensitivity=0.7), we show that increasing population-level exposure is detrimental to the study power for disease susceptibility (Supplement B).

Figure 1C shows that, combined with low exposure rate, baseline infection susceptibility given exposure also affects power, in a manner that is inversely proportional to the baseline infection rate. For example, assuming a population-level exposure rate of 20% as seen in the hardest-hit geographic regions [21], a high baseline infection rate (e.g. 90%, the purple curve in Figure 1C) diminishes the chance of detecting an associated variant with OR=0.5 and MAF=0.2 in a case-control study with 6,000 cases and 6,000 “controls”. This is because increasing baseline infection susceptibility results in more people infected *if exposed* to SARS-CoV-2, which, combined with low exposure rate, leads to a higher misclassification rate of controls and lower power (e.g. 5,268 out of 6,000 “controls” are misdassified assuming 20% population exposure and 90% baseline infection rate; Supplement A). The loss in statistical power is exacerbated by the current (fortunately) low population-level exposure to the SARS-CoV-2 infection. Recent studies have shown >80% infectivity for individuals over age 30 [15], suggesting the blue (baseline infection susceptibility 80%) or purple (baseline infection susceptibility 90%) curves in Figure 1C more closely reflect the current reality.

The lack of information on individual-level exposure, coupled with low population-level exposure rates, high infectivity and inaccurate, low-sensitivity tests, make studies designed to identify genetic variation contributing to SARS-CoV-2 infection susceptibility largely infeasible. We see that a larger study cohort, with misclassification, is not guaranteed to yield higher statistical power than a smaller dataset with well-defined cases and controls, as noted elsewhere [23]. Our simulation results indicate that careful selection of controls with a high probability of exposure (e.g. frontline workers or household members of positive cases) rather than sampling test-negative controls is a preferable strategy. Given the negative effect on power from using a low-sensitivity test, it is also preferred to include individuals who were monitored or tested multiple times in a given timeframe; such a design can potentially save costs and yield greater insights in understanding genetic susceptibility to SARS-CoV-2 infection, even with a smaller sample size.

### Identifying Host Genetic Factors of Disease Severity of COVID-19

Another major objective of host-genetic studies is to identify genetic variants associated with severity of symptoms given SARS-CoV-2 infection. Such studies may focus on severe respiratory disease [7], or alternative phenotypes identified prospectively as researchers learn more about the sequalae of infection [24]. Ideally, cases should include infected patients who have the defined phenotype and controls should be comprised of infected patients with mild or no symptoms [3,25]. One common proxy for disease severity is hospitalization [11]. However, given the time, effort and cost of recruiting and genotyping infected but non-hospitalized individuals, along with the urgent need for genetic insights, alternative designs that leverage matched controls in the general population have been proposed and implemented [7,11]. Contrary to the low population exposure rate that drives case-control misclassification in SARS-CoV-2 infection susceptibility studies, it is the low population *infection* rate (population exposure rate x baseline infection susceptibility) and the lack of information on individual-level *infection* that lead to misclassification in disease severity studies; some of the “controls” would have developed severe symptoms had they been infected with SARS-CoV-2.

Here we examine the gain in statistical power and efficiency (in terms of requiring smaller sample size) when using test-positive controls over a design that samples controls from an untested, available population sample (Figure 2). Using test-positive controls (red) yields higher power than using population-based (untested) controls (blue) and can achieve substantial savings in genotyping costs. Power, while assuming perfect test accuracy, was investigated for a common variant with moderate effect size (OR=1.7; MAF=0.2; Figure 2A); a common variant with small effect size (OR=1.3; MAF=0.2; Figure 2B); and a rare variant with large effect size (OR=5; MAF=0.01; Figure 2C). Population infection rate, effect size of the risk variant, MAF and other factors specific to the COVID-19 pandemic were varied with details provided in Supplement C.

**Figure 2:**
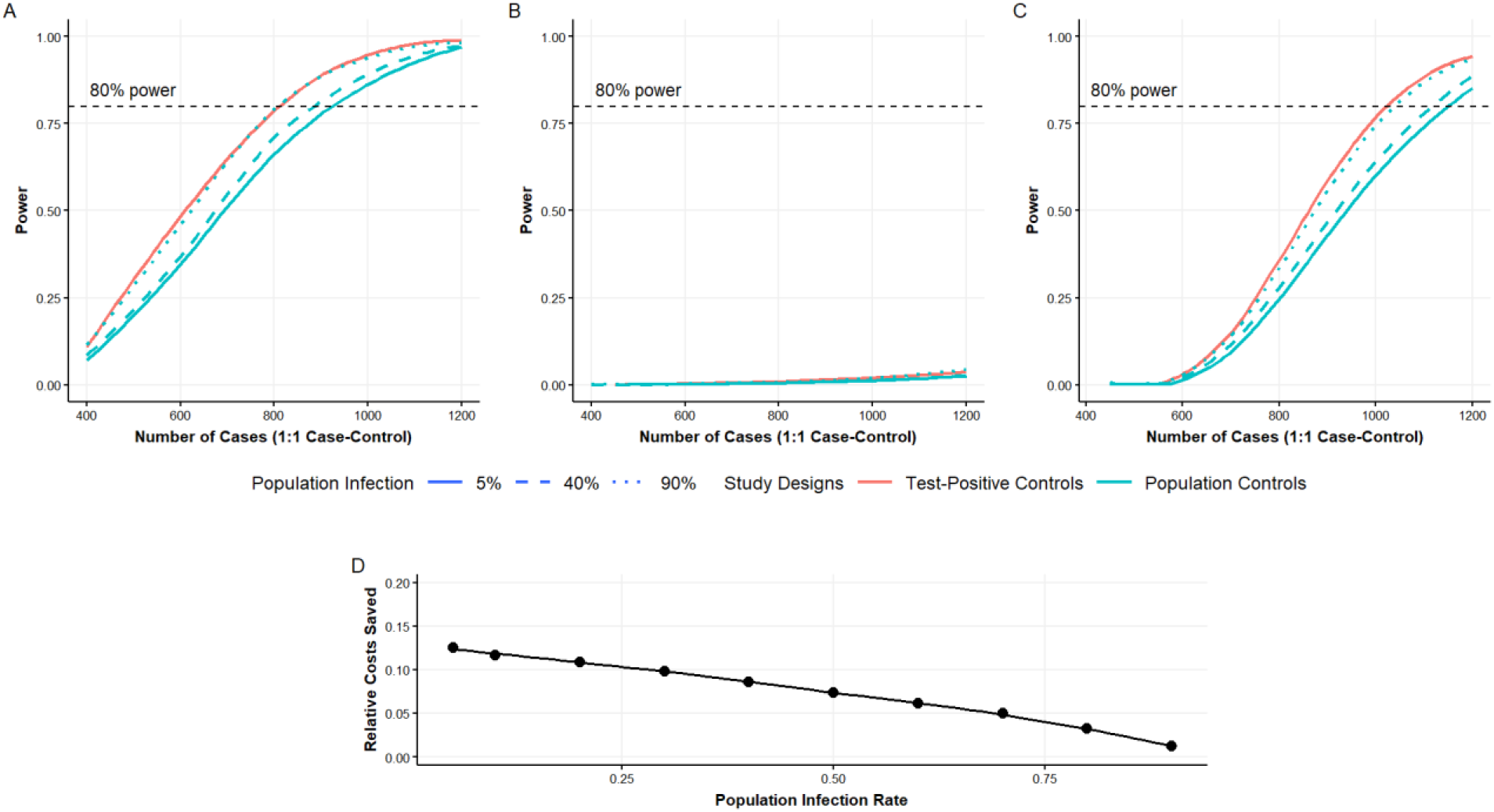
Statistical power to detect a true association between a genetic variant and COVID-19 disease severity at the genome-wide significance level (5e-8). A 1:1 case-control study design was used for all parameter settings. Only one red curve is shown since the study design uses confirmed infected individuals with mild or no symptoms as controls (test-positive controls), which is unaffected by population-level infection rates and the corresponding case-control misclassification. Effect sizes are reported on the odds ratio (OR) scale for each additional risk allele (log-additive scale). Assumes perfect test accuracy. A) Assumes a common variant with large effect size (OR=1.7, MAF=0.2). Using test-positive controls (red) yields higher power than using population-based (untested) controls (blue). High population infection rates reduce the gap between the two study designs but remains unlikely in the current phase of the pandemic. B) Detecting a common variant with moderate effect size (OR=1.3, MAF=0.2) is challenging without drastically increasing the number of participants included for either design. C) Detecting a rare variant even with a large effect size (OR=5, MAF=0.01) is more difficult at current sample size levels. Using test-positive controls without misclassification once again demonstrates higher power compared to population-based untested controls with misclassification. D) Assumes OR=1.7 and MAF=0.2. Relative reduction in sample size, 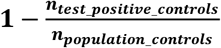, from using test-positive controls compared to population-based controls. *n_test_positive_controls_* and *n_population_controls_* refer to the number of cases (1:1 case-control ratio) needed to achieve 80% power at the genome-wide significance level (5e-8) [22]. Representative of relative reduction in sample size for other settings can be found in Supplement E.

The common (risk) variant effect sizes in Figures 2A and 2B were chosen to reflect those recently reported in genetic studies of COVID-19 [7]. Only one red curve is shown since the study design uses test-positive individuals with mild symptoms as controls, which is not influenced by population-level infection rates. Thus, there is no misclassification attributed to infection rates. As expected, the higher red curve demonstrates that severity phenotyping in the absence of misclassification in a case-control study yields higher power than using controls with misclassification from the general population. Increases in population infection rates effectively closes the power gap between the two study designs since most population-based controls would have been infected and hospitalized if necessary, thus reducing misclassification. However, as serological tests have shown low population infection rates (e.g. 5% in Spain by May 11^th^, 2020) even in the hardest-hit regions [20], we expect the gap in power demonstrated in Figure 2 to remain for the foreseeable future. This observation is most relevant for prospective studies: careful selection of individuals infected with SARS-CoV-2 as controls improves power and can reduce spending on genotyping.

The effect size of an associated risk variant can have, of course, a large impact on the power of the study design (Figure 2A vs. 2B). Detecting a common variant with moderate effect size (OR=1.3, MAF=0.2) is challenging without drastically increasing the number of participants studied. In fact, the sample size required to reach 80% power at the genome-wide significance level exceeds 4 times that required in Figure 2A (Supplement D). Similar patterns are seen when detecting a rare variant (Figure 2C) as power is lower for such variants even for a large effect size (OR=5, MAF=0.01). To achieve the same power, the smaller sample size realized by selecting test-positive rather than population controls (red curve vs blue curves in Figure 2C) once again demonstrates the potential to save significant costs in sequencing given the low population infection rates in the current phase of this pandemic.

To quantify the costs saved by using controls with confirmed infection, we defined relative reduction in sample size as 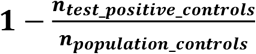. Figure 2D illustrates relative reduction in sample size with varying population infection rates when studying a common variant with a large effect size (OR=1.7, MAF=0.2); similar qualitative conclusions are gleaned as the parameters are varied (Supplement E). As expected, studies using controls with confirmed infection show the greatest benefit when disease prevalence is low. Given current levels of population infection rates at less than 10% [20], choosing controls with confirmed SARS-CoV-2 infection can save over 10% in genotyping costs. On the other hand, if genotype data for population controls are already available [9, 10, 26], ~1,200 genotyped controls are required to detect a rare variant with very large effect (OR=5.0, MAF=0.01) given (currently) low population infection rate (5%). The required sample size rises to ~3,800 for detecting a common variant with moderate effect (OR=1.3, MAF=0.2), and even more samples are needed if we allow the case-control ratio to decrease (Supplement F). Given the sheer number of population controls available internationally, this seems like a reasonable and accessible strategy. Although beyond the scope of this commentary, this strategy is limited by the challenges of confounding and host-specific factors, which are often not widely available in population-based controls, leading to difficulties in interpreting genetic association results. When sample collection is planned prospectively, these results suggest a strategy of test-positive, matched controls for optimal power and interpretation.

## Discussion

Understanding how host genetic factors contribute to variation in disease susceptibility and severity can shed light on heterogeneity in the immune response and the host-pathogen interaction, and facilitate the development of therapeutics and vaccines. Genetic studies have already been deployed in several countries to investigate SARS-CoV-2 infection susceptibility and COVID-19 disease severity. However, ongoing studies are hindered by a lack of information on SARS-CoV-2 exposure and infection at the individual level, low population exposure/infection rates and inaccurate, low-sensitivity tests. In particular, given current population exposure rates, using test-negative controls will not provide adequate power to detect variants that protect from infection when exposed to SARS-CoV-2; this strategy leads to lower study power and greater difficulty for studying infection susceptibility than disease severity. A design that focuses on, for example, frontline workers such as healthcare professionals who are at high risk of exposure and are more likely to receive repeated testing would have much greater power. For case-control studies investigating the genetics of COVID-19 disease severity, collecting those who tested positive for SARS-CoV-2 infection rather than using existing population-based controls should be prioritized whenever feasible. Household controls, which are easier to recruit and provide the added benefit of increased exposure probability, could serve as another effective selection of controls to enhance study power. Using test-positive controls also provides the added benefit of prospective phenotypic observation to allow for future genetic studies of as-of-yet unknown phenotypic sequela of this disease, even in those who are mild or asymptomatic [24]. A prospective design also facilitates future investigation into the genetics of antibody levels and vaccine responsiveness, which is of major interest in ongoing vaccine development and deployment efforts.

Contrary to the SARS-CoV-2 infection susceptibility studies, we have assumed perfect test sensitivity and specificity for the COVID-19 severity study simulations. For study designs where controls have confirmed infection, low sensitivity (false negatives) that is characteristic of RT-PCR tests used for SARS-CoV-2 infection [16] is not a major consideration. Similarily, designs that use population-based controls without leveraging information on test results, are also not affected by false negative RT-PCR tests. Previous studies have reported near-perfect specificity for RT-PCR tests [27] which leads us to conclude that imperfect test accuracy has limited impact on host-genetic studies of COVID-19 disease severity, with the exception of meeting recruitment milestones.

Thus far, we have assumed RT-PCR tests were used to identify individuals infected with SARS-CoV-2. It is worth noting that RT-PCR cannot indicate whether someone has been previously infected with SARS-CoV-2 due to its short detection period. Serology tests typically have longer detection periods, thus can be used to assess previous exposure to pathogens and contribute to a more refined selection of controls. It is also of note that, as cases and controls are currently being selected from multiple sites for practical reasons, varying test accuracies over time and location would further impact study power.

Besides exposure, there are other confounders or mediators we have not addressed in this commentary which can further reduce power, diminishing our chances to detect variants that *directly impact* COVID-19 disease severity. That is, confounding could lead to the identification of genetic variants that impact, for example, co-morbidities such as diabetes or hypertension, already known to put individuals in a higher risk category for severe disease. But these variants are likely not what would be most relevant to inform therapeutics and vaccine development. Variation associated with factors specific to the host rather than host-pathogen interactions are a major concern for case-control studies using population-based convenience controls, rather than case-control studies in captive, highly exposed populations, such as frontline workers. One example of such confounding could be the ABO locus reported in [7], where blood donors were used as a control group and type-O blood was associated with decreased risk of severe respiratory disease. Given the negative impact of ignoring important intrinsic variables on study power, pursuing stratification or matching on known intrinsic variables, especially confounders, is a desirable approach that should be considered in COVID19 case-control host genetic studies moving forward [28]. Estimates of genetic effects, measured in odds ratios, are also difficult to interpret even if the unadjusted intrinsic variables are not confounders [28]. Future studies, potentially through matching and/or stratification will also need to take into account the age distribution of cases and controls given the varying infection susceptibility and disease severity experienced in different age groups [15].

The identification of genetic variation that directly impacts infection susceptibility and disease severity would be an important step towards personalized treatment plans, risk stratification, therapeutic and vaccine development and deployment. However, limited phenotypic data and exposure/infection information significantly impact our ability to detect these variants. As the community moves ahead with the proposal and implementation of these studies, careful consideration of designs that prioritize clarity in the interpretation of the findings rather than expediency, will prove to be more beneficial and cost-effective in the long run.

## Data Availability

No data were used in this manuscript.

## Acknowledgements

Y.L. is a trainee of the CANSSI Ontario STAGE (Strategic Training for Advanced Genetic Epidemiology) program at the University of Toronto and recipient of the CANSSI COVID-19 Rapid Response Program. The senior authors represent the Genetic Epidemiology Committee of the Canadian Genomics Enterprise (CGeN) Hostseq project, which is funded by Innovation, Science and Economic Development Canada through Genome Canada.

